# Geographical structure and evolutionary trajectories of *Streptococcus pneumoniae* serotype 19F lineages in South Africa

**DOI:** 10.1101/2025.11.03.25339423

**Authors:** Antidius Laurian, Hsueh-Chien Raymond Cheng, Takudzwa Matuvhunye, Celine De Allende, Kate C Mellor, Kiran Gajanan Javkar, Heather J Zar, Mark Nicol, Stephen D Bentley, Stephanie W Lo, Felix Dube

## Abstract

**Background:** *Streptococcus pneumoniae* remains a leading cause of morbidity and mortality among children under five, causing an estimated 300,000 deaths annually worldwide. Despite widespread implementation of pneumococcal conjugate vaccines (PCVs), serotype 19F, a vaccine serotype (VT), continues to persist in several high-burden settings, including South Africa. While its persistence has been documented, the evolutionary and spatial mechanisms underpinning continued circulation remain poorly understood. We investigated the evolutionary history and fine-scale spatial dynamics of predominant serotype 19F lineages within the Drakenstein Child Health Study (DCHS) to better understand the processes maintaining persistence in a highly vaccinated population.

**Methods:** We analysed whole-genome sequences from 617 serotype 19F carriage isolates collected from infants enrolled in the DCHS, Cape Town, South Africa (2012–2017). To investigate long-term evolutionary dynamics, local genomes were combined with publicly available South African serotype 19F genomes from the Global Pneumococcal Sequencing (GPS) project. Bayesian molecular dating and complementary spatial genetic analyses were used to investigate lineage evolution and fine-scale transmission dynamics.

**Results:** Three Global Pneumococcal Sequence Clusters (GPSC205, GPSC1, and GPSC21) accounted for 98% of all serotype 19F isolates. GPSC1 and GPSC21 exhibited significant temporal signal with the evolutionary rate of 10.3 and 5.6 substitutions per year, respectively, whereas GPSC205 showed no measurable molecular clock signal. Relative risk (RR) analyses demonstrated significant spatial clustering of isolates belonging to the same lineage within 1 km (RR = 1.32 (95% CI: 1.09–1.80)). However, genetic similarity showed a weak relationship with geographic proximity (r = 0.063, p = 0.0004), and geographic distances between isolate pairs remained largely unchanged across divergence times, indicating extensive genetic mixing across the study area.

**Conclusions:** Persistence of pneumococcal serotype 19F appears to be driven by the long-term co-circulation of successful lineages rather than sustained expansion of geographically restricted clones. Integrating evolutionary and spatial genomic analyses provides a more comprehensive understanding of VTs’ persistence and may strengthen genomic surveillance in highly vaccinated populations.

## 1.0 INTRODUCTION

*Streptococcus pneumoniae* (pneumococcus) is a Gram-positive bacterium that asymptomatically colonises the upper respiratory tract (URT), particularly the nasopharynx. While often carried without symptoms, colonisation is a prerequisite for transmission and the development of pneumococcal disease. Pneumococcal infections account for an estimated 300,000 deaths annually in children under five years of age (Al-Jumaili et al., 2023). The burden is highest in low- and middle-income countries, where social and environmental factors such as poverty, overcrowding, and high HIV prevalence amplify susceptibility.

Though typically opportunistic, pneumococci can cause invasive pneumococcal disease (IPD), including bacteraemia, meningitis, and sepsis, as well as lower respiratory tract infections such as pneumonia and bronchitis (Cedrone et al., 2023). Colonisation is extremely common in children, approaching 95% within the first year of life (Dube et al., 2018), while adult carriage rarely exceeds 10% (Weiser et al., 2018). Transmission usually occurs through close contact with respiratory secretions from carriers or infected individuals (Zivich et al., 2018). Furthermore, children in daycare or large households often act as vectors seeding infection within families (Melegaro et al., 2004).

The pneumococcal capsule is the primary virulence determinant, conferring resistance to phagocytosis and complement-mediated killing (Hyams et al., 2010; Loughran et al., 2019; Mitchell & Mitchell, 2010). Serological and structural variation in the capsule forms the basis for classification into over 108 serotypes. Importantly, capsule diversity influences disease potential, transmission fitness, and vaccine effectiveness. Whole-genome sequencing (WGS) has enabled further lineage-level resolution through Global Pneumococcal Sequencing Clusters (GPSCs), identified using genome-wide approaches such as PopPUNK (Lees et al., 2019). Currently, there are more than 1000 pneumococcal GPSCs. GPSCs enable analysis of global genomic datasets, providing insights into capsular switching, the spread of antimicrobial resistance (AMR), and lineage replacement under vaccine pressure (collection, 2026; Lo et al., 2019; Von Mollendorf et al., 2017).

Pneumococcal serotype 19F has proven unusually persistent despite the widespread implementation of pneumococcal conjugate vaccines (PCVs) which include this serotype (Javaid et al., 2022; Mokaya et al., 2023; Usuf et al., 2014). Its continued circulation highlights the complex interplay between bacterial adaptability, host immunity, antibiotic exposure, and population dynamics. Compared to other vaccine serotypes, 19F exhibits reduced immunogenicity, higher carriage density, and frequent association with multidrug resistance (MDR) (Osei et al., 2025; Shi et al., 2024). These features contribute not only to its survival but also to clinical challenges, including treatment failures (Feemster et al., 2023).

In South Africa, PCV7 was introduced in 2009 before being replaced by PCV13 in 2011. While these vaccines dramatically reduced disease caused by vaccine serotypes, residual carriage of serotype 19F has persisted in communities such as the Drakenstein region of the Western Cape, where vaccine uptake exceeds 98% (Dube et al., 2018). The persistence of serotype 19F in highly vaccinated populations represents a paradox of modern pneumococcal control, likely influenced by its stable capsule, lineage-specific acquisition of antimicrobial resistance, and environmental conditions that facilitate transmission (von Gottberg et al., 2024).

Although previous studies have demonstrated the continued circulation of serotype 19F (Agudelo et al., 2021; Patel et al., 2022; Skosana et al., 2021), the evolutionary and spatial processes underlying its persistence remain poorly understood. In particular, it is unclear whether persistence is driven by the long-term maintenance of successful lineages, continual local transmission of closely related strains, or repeated introduction and co-circulation of genetically diverse members of the same lineage. Furthermore, the extent to which these lineages exhibit measurable temporal signal, accumulate genetic divergence over time, and disperse across geographic space has not been comprehensively investigated.

Addressing these questions requires a high-resolution investigation of pathogen transmission. Pathogen migration and spread across communities are fundamental components of infectious disease dynamics (Kozakiewicz et al., 2018). Genomic epidemiology provides powerful tools to investigate these processes by integrating pathogen genomes with epidemiological and spatial data. Linking genetic variation to temporal and geographic information enables reconstruction of transmission pathways and characterization of lineage dynamics under vaccine- and antibiotic-mediated selective pressures (Lo et al., 2022; Pronyk et al., 2023). Relative risk (RR) frameworks, for example, have been successfully applied to quantify spatial clustering of genetically related strains and to infer transmission patterns within and between communities (Belman et al., 2024; Cheng et al., 2024).

In this study, we investigated both the evolutionary and spatial persistence of serotype 19F. First, we examined the evolutionary dynamics of the predominant lineages by assessing their temporal signal, estimating evolutionary rates, and investigating the relationship between divergence time and geographic distance using a combined dataset of local isolates from the Drakenstein and publicly available genomes from the Global Pneumococcal Sequencing (GPS) project. Second, we investigated spatial persistence within the Drakenstein cohort by characterising the local distribution of serotype 19F lineages and quantifying their spatial clustering to determine whether persistence is primarily associated with localized transmission of closely related isolates or with the co-circulation of genetically diverse members of the same lineage.

The Drakenstein cohort provides a uniquely detailed carriage dataset, combining dense longitudinal sampling, high vaccine coverage, and well-characterized household locations. Leveraging these data together with global genomic data, this study provides a comprehensive assessment of the evolutionary history and fine-scale spatial epidemiology of serotype 19F, offering new insights into the mechanisms that enable this vaccine serotype to persist despite sustained vaccination.

## 2.0 METHODS

### 2.1 Study design and participants

Details of the study area, design, and participants have been previously described (Manenzhe et al., 2019). Briefly, the study was conducted at two primary healthcare clinics, TC Newman and Mbekweni, located approximately 2 km apart in the Drakenstein region of the Western Cape, South Africa. This is a nested study within the broader Drakenstein Child Health Study (DCHS), a population-based, longitudinal, and prospective birth cohort based in the Western Cape Province, South Africa. Ethical approval for the study was obtained from the Faculty of Health Sciences, Human Research Ethics Committee (HREC) of the University of Cape Town, South Africa (401/2009).

Procedures for nasopharyngeal (NP) sample collection, transport, culture, and storage have been described elsewhere (Dube et al., 2018). In brief, all infants received routine immunization, including PCV13 as part of the national immunization program, in a 2+1 dosing schedule at 6 weeks, 14 weeks, and 9 months of age. NP samples were collected every 2 weeks during the first year of life, and subsequently every 6 months until five years of age. Between 26 June 2012 and 25 April 2018, 19219 NP samples were obtained from 1,090 infants born to 1,013 enrolled mothers. Procedures for pneumococcal isolation, DNA extraction, sequencing, and genome assembly have been described previously (Mackenzie et al., 2016). Briefly, DNA was paired-end sequenced on an Illumina HiSeq platform at the Wellcome Sanger Institute. All isolates were processed through the Global Pneumococcal Sequencing (GPS) pipeline for quality control (QC), which included read trimming, contamination screening, and assembly quality assessment (Hung et al., 2025). The serotype 19F subset analysed in this study comprised exclusively QC-passed genomes. Serotypes, GPSCs, Clonal Complexes (CCs), and Sequence Types (STs) were inferred using PathogenWatch v23.5.0 (https://pathogen.watch/) (Gladstone et al., 2020).

### 2.2 Data curation and inclusion criteria

A total of 617 serotype 19F isolates were available from the Drakenstein cohort. Because several participants contributed multiple isolates collected at different time points, we retained a single representative isolate per participant within each GPSC for all inferential spatial and evolutionary analyses to avoid pseudoreplication arising from repeated sampling. The resulting dataset was further restricted to the three dominant GPSCs (GPSC1, GPSC21, and GPSC205), each represented by at least 30 isolates, yielding a final dataset of 239 genomes **(Figure 1)**. Relative risk, Mantel, divergence time–geographic distance, and spatial homogenization analyses were all performed using this dataset. The exclusion of the remaining rare GPSCs (GPSC5, GPSC90, and GPSC192) did not materially alter the relative risk estimates but improved consistency across analyses and ensured robust statistical inference.

**Figure 1.**
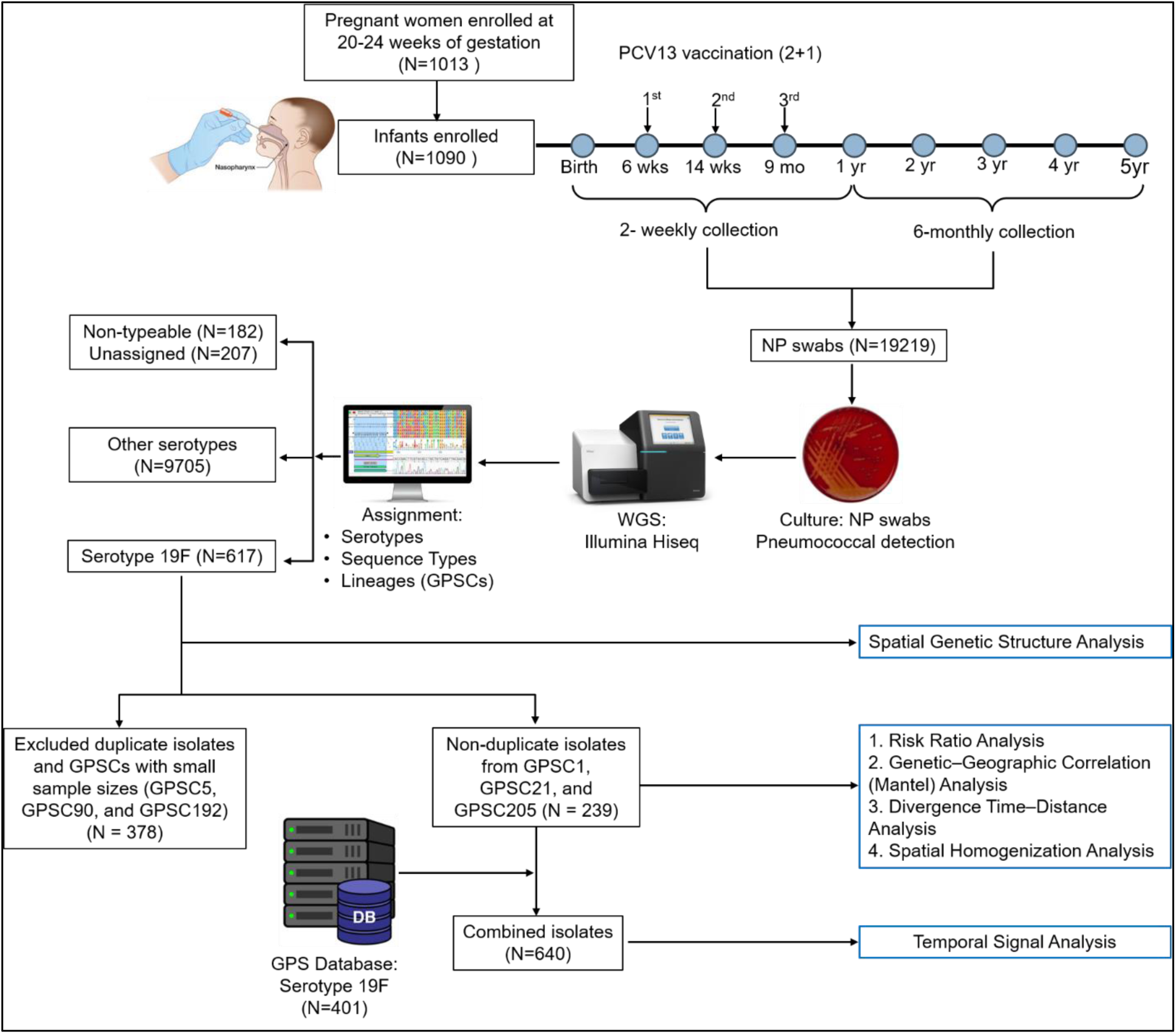
Workflow of participant enrolment, sample processing, dataset curation, and downstream analyses. Children enrolled in the Drakenstein birth cohort underwent scheduled nasopharyngeal sampling throughout early childhood and received pneumococcal conjugate vaccine (PCV) according to the study schedule. Pneumococcal isolates were cultured, whole-genome sequenced, and processed through genomic analysis pipelines. A total of 617 local serotype 19F isolates were initially available. For inferential spatial and evolutionary analyses, repeated isolates from the same participant were removed by retaining one representative isolate per participant within each Global Pneumococcal Sequencing Cluster (GPSC), and analyses were restricted to the dominant lineages (GPSC1, GPSC21, and GPSC205), yielding 239 local isolates. These isolates were used for relative risk analysis, genetic–geographic correlation analysis, divergence time–geographic distance analysis, and spatial homogenization analysis. To evaluate the temporal signal, the 239 local isolates were combined with 401 serotype 19F genomes retrieved from the Global Pneumococcal Sequencing (GPS) database, resulting in a dataset of 640 isolates spanning approximately two decades for time-scaled phylogenetic and temporal evolutionary analyses.

Pneumococcal lineages were defined using the GPSC framework, which classifies *Streptococcus pneumoniae* based on whole-genome similarity. This standardized nomenclature captures genome-wide population structure and facilitates comparisons with international datasets.

To evaluate the temporal signal, the 239 local isolates were supplemented with 401 serotype 19F genomes obtained from the GPS database, resulting in a combined dataset of 640 isolates. Because the local Drakenstein isolates were collected over a relatively short period (2012–2017), the additional genomes extended the temporal sampling window to approximately two decades (2001–2020), thereby increasing the power to detect temporal signal and estimate evolutionary rates. Only serotype 19F isolates from South Africa, belonging to GPSC1 (N=160), GPSC21 (N=224), and GPSC205 (N=17), were included. The GPS dataset was accessed on 9 March 2026, and the most recent eligible genome had been collected in 2020.

### 2.3 Sequence alignment

For each dominant GPSC, a reference genome was selected from the GPS project: *S pneumoniae* Taiwan 19F-14 for GPSC1 (NCBI accession number CP000921), and strain ZA_GPS_SP437 for GPSC21 (NCBI accession number GCA_901218315). The reference genome can also be downloaded from the GPS project (https://figshare.com/articles/online_resource/GPSC_references/12783260?file=44894077).

Last updated 06 March 2024. The list, however, did not include the reference for GPSC205. Therefore, we obtained a genome from the GPS project database that had the lowest number of contigs, no apparent contamination, and had a ‘PASS PLUS’ status; Sanger sample ID: 2245STDY5883326, public name: GPS_ZA_1822, accessed on 02 April 2026 (Gladstone et al., 2020). Sequences were then aligned to the corresponding references using the split *k*-mer tool (SKA2) v0.4.0 (Derelle et al., 2024).

### 2.4 Generating a recombination-free tree

Recombination sites were masked by Gubbins v3.4.2, which identifies loci containing elevated densities of base substitutions that are marked as recombination while concurrently constructing a phylogeny based on the putative point mutations outside these regions. Phylogenetic trees were generated by RAxML v8.2.12 and a general time-reversible (GTR) model (Croucher et al., 2015; Stamatakis, 2014) allowing for different rates of substitution between each nucleotide pair while maintaining reversibility (Hung et al., 2025). These computations were performed in the Ilifu cloud computing facility (www.ilifu.ac.za).

### 2.5 Estimation of the temporal signal of predominant serotype 19F lineages

To determine whether the predominant serotype 19F lineages accumulated genetic variation in a clock-like manner over time, we estimated their temporal signal using the combined dataset of 640 isolates (239 Drakenstein isolates and 401 serotype 19F isolates from the GPS project). The mean evolutionary rates and ancestral dates were inferred using BactDating under an additive uncorrelated relaxed clock (ARC) model (Didelot et al., 2018; Didelot et al., 2021). BactDating applies a Bayesian framework to infer evolutionary timescales from dated phylogenetic trees.

For each GPSC, sampling years were extracted from the metadata and matched to the corresponding phylogenetic tip labels. Root-to-tip genetic distances (substitutions per site) were then calculated and regressed against sampling years to evaluate the strength of the temporal signal and estimate lineage-specific evolutionary rates.

### 2.6 Estimation of the relationship between spatial distance and divergence time

To investigate how spatial separation changes with evolutionary divergence, we examined the relationship between geographic distance and divergence time among isolate pairs. Pairwise divergence times were estimated using the posterior distribution of time-scaled phylogenies generated by BactDating, while geographic distances were calculated from household coordinates.

Representative phylogenetic trees were sampled at regular intervals from the Markov chain Monte Carlo (MCMC) posterior. For each sampled tree, mean pairwise divergence times were calculated and grouped into predefined divergence time windows. To account for uncertainty in these estimates, bootstrap resampling was performed within each divergence window. Mean geographic distances and corresponding 95% confidence intervals were then calculated for each lineage.

### 2.7 Spatial distribution

We first visualized the spatial distribution of serotype 19F lineages across the Drakenstein study area. Latitude and longitude coordinates for all 617 isolates were mapped and coloured according to their corresponding GPSC. Maps were generated using *ggplot2* (Wickham, 2016) package in R v4.2.2 (Posit team, 2025).

### 2.8 Assessing spatial clustering using the relative risk ratio

While spatial maps illustrate the geographic distribution of lineages, they do not quantify whether isolates belonging to the same lineage cluster together in space. Therefore, we adapted the relative risk (RR) framework (Belman et al., 2023; Cheng et al., 2024) to quantify the extent of spatial clustering among serotype 19F lineages.

Specifically, the RR compares the probability that two isolates belong to the same GPSC within a given spatial distance with the corresponding probability among geographically distant isolate pairs (reference distance). An RR greater than one indicates that isolates belonging to the same lineage are more likely to occur within that spatial window than expected from the background distribution.

For each spatial window, we calculated the proportion of isolate pairs belonging to the same GPSC among all isolate pairs within that distance. This probability was then divided by the corresponding probability calculated for the reference distance window (5–9 km).

The framework described above can be briefly summarized as follows.

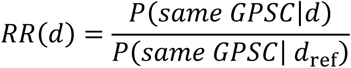

Where *RR*(*d*) is the relative risk of finding two isolates of the same GPSC at a given spatial distance *d*, compared to the baseline, *d*_ref_ (reference distance window (5–9 km)), and *P* is the probability that a pair of isolates belongs to a given group.

The reference distance window (5–9 km) was selected based on the empirical distribution of pairwise geographic distances. Approximately 75% of isolate pairs were separated by ≤5.3 km; therefore, distances ≥5 km were considered representative of geographically separated isolate pairs. Extending the upper limit to 9 km increased the number of pairwise comparisons contributing to the baseline estimate while ensuring that the reference represented the most spatially separated isolate pairs available in the dataset.

### 2.9 Spatial homogenization over evolutionary time

To estimate the timescale over which serotype 19F lineages become geographically dispersed throughout the Drakenstein study area, we conducted a rolling-window analysis across divergence time intervals (0–2, 2–4, 4–6, 6–8, 8–10, 10–15, 15–25, and 25–35 years). Relative risk ratios were calculated within each divergence time window using both household-level geographic distances and regional classifications. These divergence intervals were selected to encompass the full range of pairwise divergence times inferred across the three dominant GPSCs.

### 2.10 Relationship between genetic and geographic distances

Finally, we assessed whether geographic proximity corresponded to greater genetic similarity among isolates using a Mantel test with Spearman’s rank correlation.

Genetic distances were represented by a patristic distance matrix derived from the phylogenetic trees, whereas geographic distances were calculated from household coordinates using the Haversine formula. Statistical significance was assessed using 999 permutations. Scatter plots with fitted regression lines were generated to visualize the relationship between genetic and geographic distances.

## 3.0 RESULTS

### 3.1 Participant characteristics

Of the 1,090 infants enrolled in this study, 617 serotype 19F isolates were obtained from 242 infants, forming the basis of the current analysis. The demographic, socioeconomic, and clinical characteristics of the participants are summarized in **Table 1**, excluding 75 infants (13 from the “serotype 19F” group and 62 from the “other serotypes” group) with missing data.

**Table 1:** Participant characteristics in the “serotype 19F” and the “other serotypes” groups of the Drakenstein birth cohort.

| Variable | Category | Serotype 19F (N=229) |  | Other serotypes (N=786) |  |
| --- | --- | --- | --- | --- | --- |
|  |  | n (%) | Missing data (n) | n (%) | Missing data (n) |
| Sociodemographic characteristics |  |  |  |  |  |
| Site | Mbekweni | 127 (55.5) |  | 441 (56.1) |  |
|  | TC Newman | 102 (44.5) |  | 345 (43.9) |  |
| Sex | Male | 121 (52.8) |  | 399 (50.8) |  |
|  | Female | 108 (47.2) |  | 387 (49.2) |  |
| Ethnicity | Black African | 129 (56.3) |  | 438 (55.8) | 1 |
|  | Mixed Ancestry | 100 (43.7) |  | 347 (44.2) |  |
| Birth season | Autumn | 55 (24.0) |  | 199 (25.3) |  |
|  | Spring | 53 (23.1) |  | 180 (22.9) |  |
|  | Summer | 55 (24.0) |  | 200 (25.4) |  |
|  | Winter | 66 (28.8) |  | 207 (26.3) |  |
| Delivery type | Vaginal | 182 (79.8) | 1 | 629 (80.3) | 3 |
|  | Caesarean | 46 (20.2) |  | 154 (19.7) |  |
| HIV status at birth | HIV exposed | 51 (22.3) |  | 178 (22.6) |  |
|  | HIV unexposed | 178 (77.7) |  | 608 (77.4) |  |
| Birth term | Early term | 22 (9.6) | 1 | 136 (17.3) |  |
|  | Full term | 206 (90.4) |  | 650(82.7) |  |
| Birth weight | NBW (≥2.5kg) | 200 (87.7) | 1 | 661 (85.1) | 9 |
|  | LBW (<2.5kg) | 28 (12.3) |  | 116 (14.9) |  |
| Exclusive breastfeeding | Yes | 139 (61.2) | 2 | 427 (54.8) | 7 |
|  | No | 88 (38.8) |  | 352 (45.2) |  |
| Number of children in household | Small (≤1 child) | 162 (72.0) | 1 | 594 (76.6) |  |
|  | Medium (2–3 children) | 63 (28.0) |  | 181 (23.4) |  |
|  | Large (≥4 children) | 3 (1.3) |  | 11 (1.6) |  |
| Number in household | Small (≤3 members) | 65 (28.1) | 1 | 167 (24.0) | 100 |
|  | Medium (4–6 members) | 115 (49.8) |  | 352 (50.5) |  |
|  | Large (≥7 members) | 48 (20.8) |  | 167 (24.0) |  |
| Clinical data |  |  |  |  |  |
| PCV at 6 weeks | Yes | 225 (99.6) | 3 | 763 (99.6) | 20 |
|  | No | 1 (0.4) |  | 3 (0.4) |  |
| PCV at 14 weeks | Yes | 221 (99.1) | 6 | 741 (99.6) | 42 |
|  | No | 2 (0.9) |  | 3 (0.4) |  |
| PCV at 9 months | Yes | 209 (98.6) | 17 | 707 (98.7) | 70 |
|  | No | 3 (1.4) |  | 9 (1.3) |  |
| Current antibiotic use | Yes | 3 (1.7) | 50 | 11 (1.8) | 185 |
|  | No | 176 (98.3) |  | 590 (98.2) |  |
| Antibiotic use past week* | Yes | 2 (1.1) | 51 | 4 (0.7) | 196 |
|  | No | 176 (98.9) |  | 586 (99.3) |  |

75 Participants were missing all the presented data except the serotype category, hence were excluded from the table.
Abbreviations: N, total number of isolates; n, number of isolates.

Among children carrying serotype 19F, most were born at full term (90.4%, 206/228) and delivered vaginally (79.8%, 182/228). Births were distributed relatively evenly across the four seasonal categories, showing no marked seasonal predominance. Household crowding appeared moderate overall, as most children lived in homes with few to moderate numbers of household members and siblings. PCV13 vaccination coverage was notably high, with more than 98% of participants having received all three recommended doses.

### 3.2 Serotype 19F prevalence and population structure

Serotype 19F (6.4%, 617/9705) was the most prevalent PCV13-included serotype in the Drakenstein cohort (**Figure 2**).

**Figure 2:**
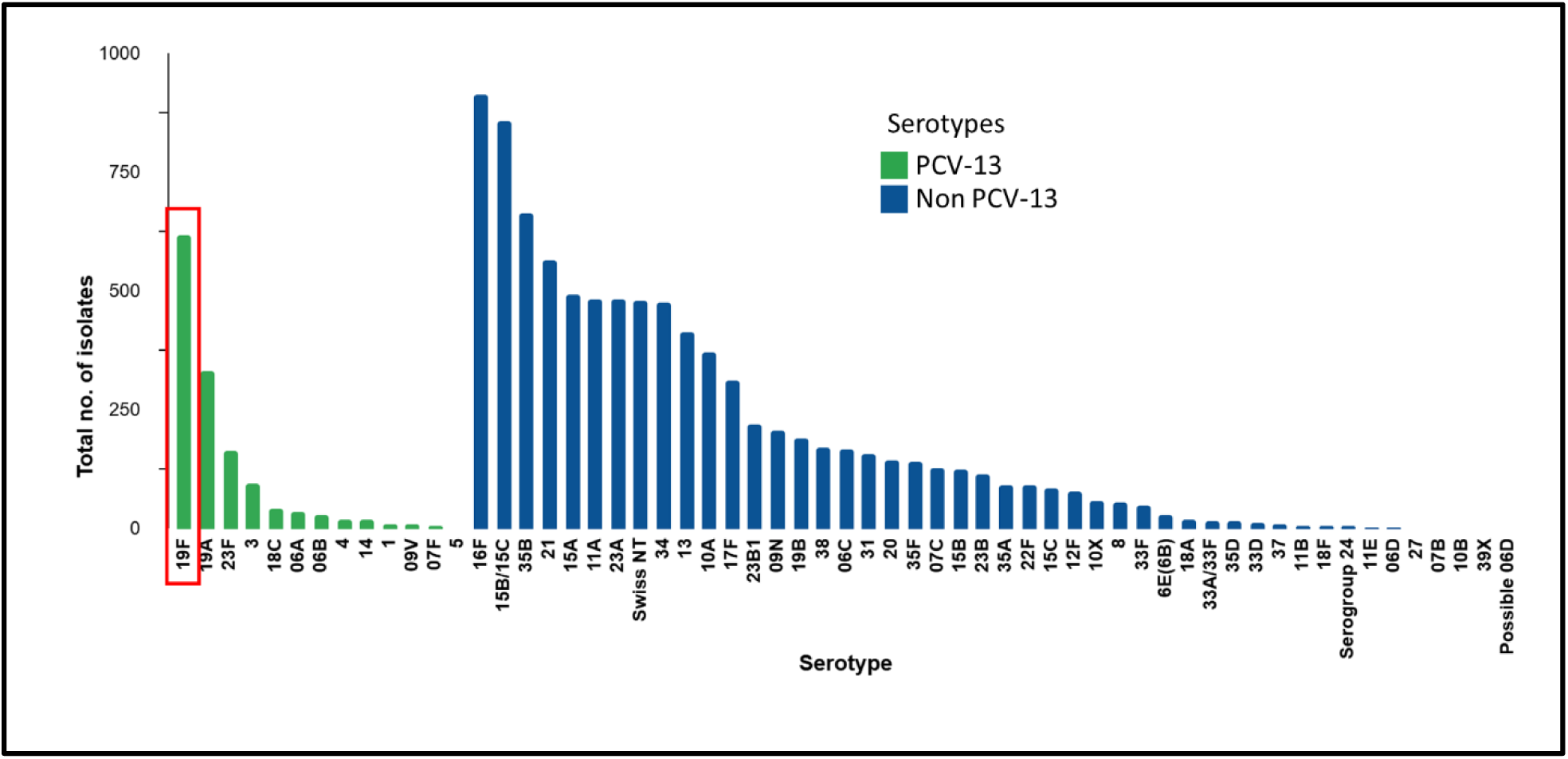
Distribution of pneumococcal serotypes in the Drakenstein cohort, 2012–2018. Bars represent the total number of isolates identified per serotype. Serotypes included in the 13-valent pneumococcal conjugate vaccine (PCV13) are shown in green, while non-PCV13 serotypes are shown in blue. Among the vaccine serotypes, 19F was the most prevalent (marked with the red rectangle). Among the non-vaccine serotypes, 16F was the most identified.

Six GPSCs were identified, with the majority belonging to GPSC205 (50.1%, 309/617), GPSC1 (35.7%, 220/617), and GPSC21 (12.3%, 76/617). These three GPSCs accounted for over 98% of all serotype 19F isolates. Additionally, 27 STs were identified with ST10823 (42.5%, 262/617), ST271 (21.6%, 133/617), and ST347 (7.9%, 49/617) being the most prevalent. These STs were strongly associated with specific GPSCs: ST10823 with GPSC205, ST271 with GPSC1, and ST347 with GPSC21.

There were three major CCs: CC2067 (50.1%, 309/617), CC271 (33.2%, 205/617), and CC347 (9.6%, 59/617). The remaining isolates were distributed among nine singleton STs associated with five different GPSCs and none of which were assigned to GPSC205 **(Table 2)**.

**Table 2:** Characteristics of serotype 19F isolates from the Drakenstein cohort.

| Variable | Proportion (N = 617)<br>n (%) | Variable | Proportion (N = 617)<br>n (%) |
| --- | --- | --- | --- |
| **Lineage (GPSC)** |  | **Clonal Complex (CC)** |  |
| 1 | 220 (35.7) | 2067 | 309 (50.1) |
| 5 | 8 (1.3) | 271 | 205 (33.2) |
| 21 | 76 (12.3) | 347 | 59 (9.6) |
| 90 | 2 (0.3) | **Singletons** |  |
| 192 | 2 (0.3) | 420 | 12 (1.9) |
| 205 | 309 (50.1) | 17666 | 12 (1.9) |
| **Sequence Type (ST)** |  | 172 | 8 (1.3) |
| 10823 | 262 (42.5) | 17566 | 5 (0.8) |
| 271 | 133 (21.6) | 10740 | 2 (0.3) |
| 347 | 49 (7.9) | 17648 | 2 (0.3) |
| 12134 | 37 (6.0) | 8328 | 1 (0.2) |
| 2067 | 32 (5.2) | 17619 | 1 (0.2) |
| 8678 | 16 (2.6) | 17613 | 1 (0.2) |
| 12535 | 14 (2.3) | **Year of isolation** |  |
| 420 | 12 (1.9) | 2012 | 1 (0.2) |
| 17666 | 12 (1.9) | 2013 | 101 (16.4) |
| 172 | 8 (1.3) | 2014 | 120 (19.4) |
| 11696 | 8 (1.3) | 2015 | 245 (39.7) |
| 15351 | 6 (1.0) | 2016 | 124 (20.1) |
| 8671 | 5 (0.8) | 2017 | 26 (4.2) |
| 17566 | 5 (0.8) |  |  |
| 15373 | 3 (0.5) |  |  |
| 10740 | 2 (0.3) |  |  |
| 11238 | 2 (0.3) |  |  |
| 17648 | 2 (0.3) |  |  |
| 6277 | 1 (0.2) |  |  |
| 8328 | 1 (0.2) |  |  |
| 10598 | 1 (0.2) |  |  |
| 15345 | 1 (0.2) |  |  |
| 17612 | 1 (0.2) |  |  |
| 17613 | 1 (0.2) |  |  |
| 17619 | 1 (0.2) |  |  |
| 17649 | 1 (0.2) |  |  |
| 17653 | 1 (0.2) |  |  |

358 Abbreviations: N, total number of isolates; n, number of isolates; GPSC, global pneumococcal sequencing clusters.

### 3.3 Temporal evolutionary dynamics of predominant serotype 19F lineages

The three GPSCs were identified across all nine South African provinces, although their geographical distributions differed **(Figure 3)**. Gauteng Province contained the highest number of isolates and represented all three GPSCs. GPSC205 showed the most restricted geographical distribution, occurring only in the Western Cape, Northern Cape, and Eastern Cape provinces, whereas GPSC1 and GPSC21 were more widely distributed. GPSC21 was the only lineage detected in Limpopo Province.

**Figure 3.**
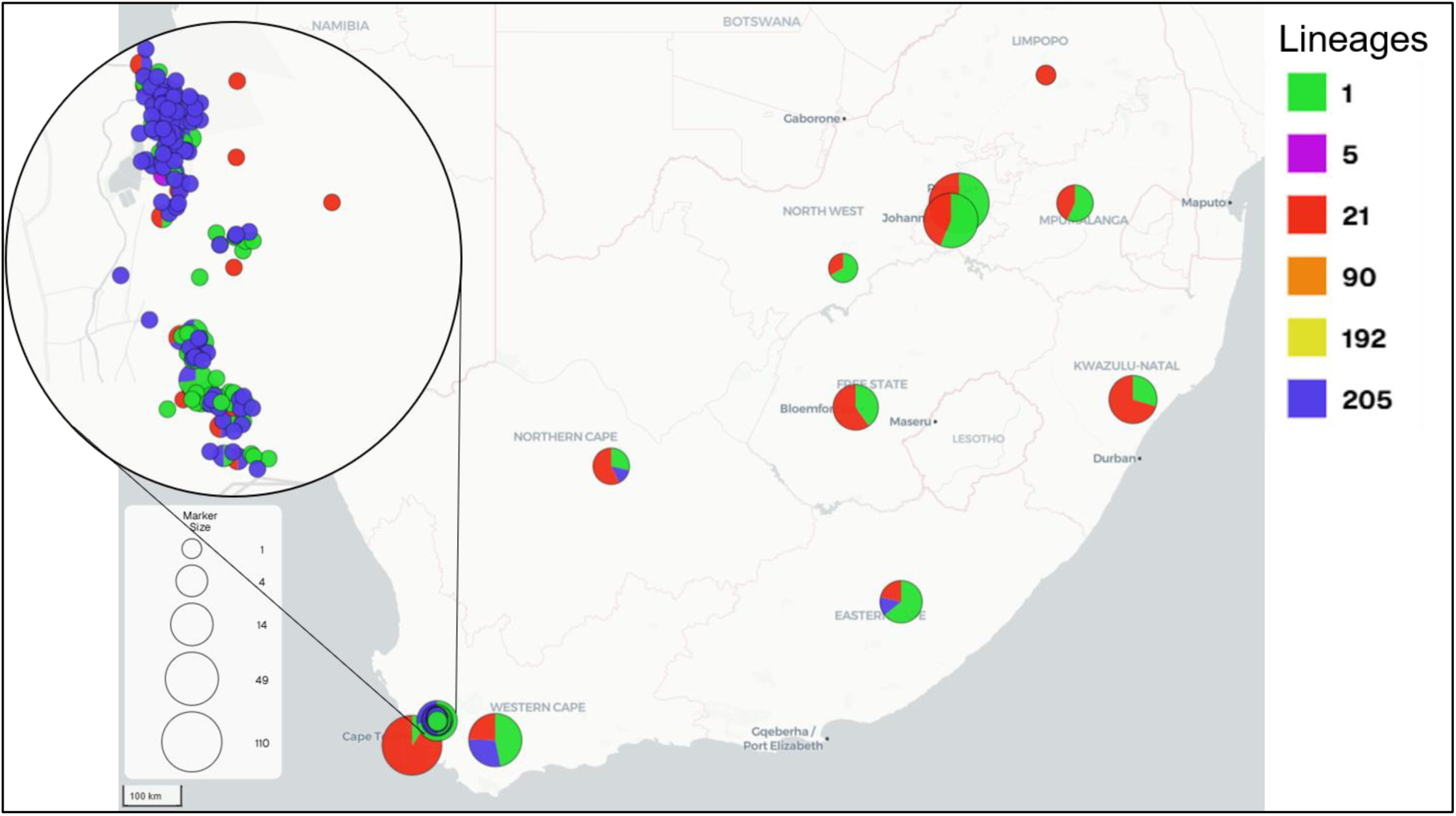
Geographic distribution of serotype 19F lineages across South Africa. Pie charts represent the proportional distribution of the lineages within each province. Pie chart size is proportional to the number of isolates sampled from each province. The inset shows the geographic distribution of serotype 19F isolates from the Drakenstein, with each point representing an individual isolate coloured according to the lineage.

Phylogenetic reconstruction revealed multiple lineage-specific clusters composed exclusively of isolates collected during the post-PCV period across GPSC1, GPSC21, and GPSC205. Similarly, several distinct clusters consisted entirely of isolates from the local Drakenstein cohort, while others contained a mixture of local and global isolates, demonstrating varying degrees of genetic relatedness between South African and internationally sourced genomes **(Figure S1)**.

The root-to-tip regression identified statistically significant temporal signals for GPSC1 and GPSC21, with estimated evolutionary rates of 10.3 and 5.6 substitutions per year, respectively (GPSC1: R² = 0.04, p = 0.0012; GPSC21: R² = 0.06, p < 0.0001). Although both lineages exhibited significant temporal structure, the relationships between genetic divergence and sampling time were weak. In contrast, GPSC205 showed no evidence of a temporal signal, with a negative regression slope (−1.05 substitutions per year), a very weak relationship between genetic divergence and sampling time (R² = 0.02), and a non-significant permutation test (p = 0.945).

We then analyzed the relationship between the divergence time and geographical distance. We found that for GPSC21 and GPSC1, the geographical distance between pairs of isolates increased with divergence time, reaching approximately 500 km for isolate pairs that had diverged for ≥8 years (95% CI: 354.29–522.48 km) and ≥14 years (95% CI: 418.16–566.01 km), respectively. In contrast, the geographical distance among GPSC205 isolates remained relatively stable across all divergence times (95% CI: 2.51–14.31 km) **(Figure 4)**.

**Figure 4:**
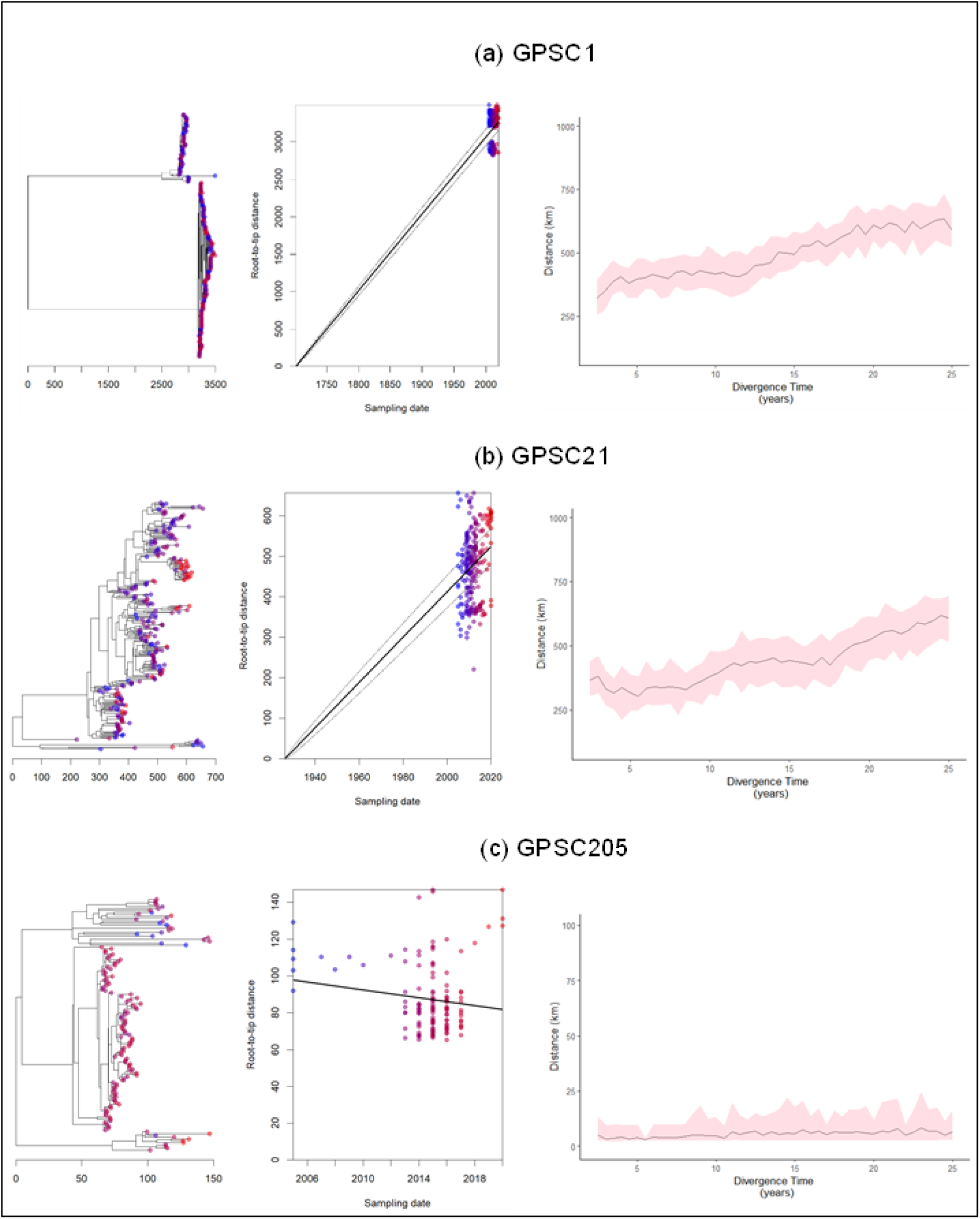
Temporal signal and genetic diversity of serotype 19F lineages. Root-to-tip regression plots and corresponding phylogenetic trees showing the evolutionary signal over time. In all panels, individual points are coloured by sampling year, with a gradient from blue (earliest samples) to red (most recent samples). The black line in each regression plot represents the best-fit line of root-to-tip genetic distance versus sampling date. To the right are plots showing the relationship between pairwise cumulative divergence time and mean geographical distance with 95 % credible interval (red). GPSC, Global Pneumococcal Sequencing Cluster.

### 3.4 Spatial genetic structure of predominant serotype 19F lineages in the Drakenstein

Having established the temporal evolutionary dynamics of the predominant serotype 19F lineages, we next investigated whether their persistence within the Drakenstein cohort was associated with fine-scale spatial genetic structure.

#### 3.4.1 Spatial distribution of predominant lineages

The spatial distribution of serotype 19F isolates revealed two major geographical clusters across the Drakenstein study area **(Figure 5a)**. GPSC1 (red) and GPSC205 (pink) were widely distributed throughout the study area, whereas GPSC21 showed a more localized distribution.

**Figure 5:**
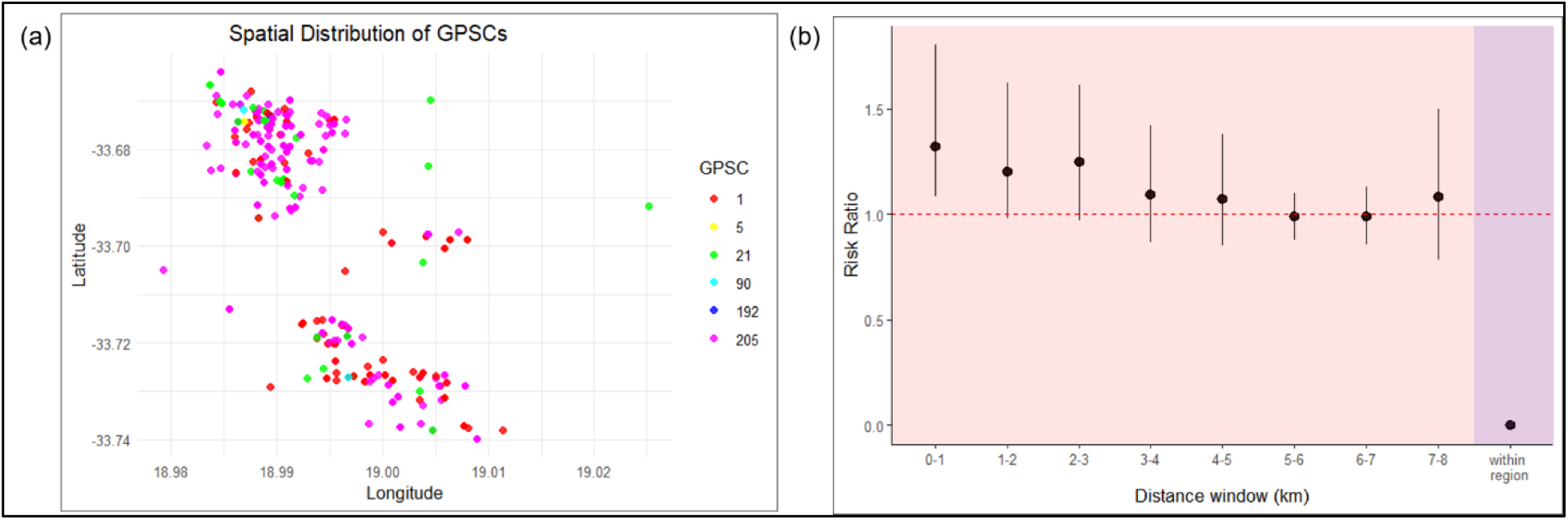
The spatial distribution of serotype 19F lineages in the Drakenstein. (a) A scatter plot showing the spatial distribution of isolates based on their geographical coordinates. Each point represents an isolate, colour-coded by its GPSC. (b) Relative risk ratio estimates for pneumococcal lineages distribution across distance windows. The red dashed line indicates a relative risk of 1 (no difference in risk). Pink shading indicates distance-based comparisons, while purple shading indicates within-region comparisons. Solid black dots and lines represent the 2.5, 50, and 97.5 percentiles of the confidence intervals. GPSC, Global Pneumococcal Sequencing Cluster.

#### 3.4.2 Relative risk

Although the spatial maps suggest clustering, they do not quantify whether isolates belonging to the same lineage occur closer together than expected by chance.

Overall, isolate pairs belonging to the same GPSC were 1.32 times (95% CI: 1.09–1.80) more likely to occur within 1 km than geographically distant isolate pairs (≥5 km reference distance). The RR progressively declined with increasing spatial separation, approaching unity at distances of 5–6 km **(Figure 5b)**.

#### 3.4.3 Relationship between genetic and geographic distance

Despite the observed spatial clustering at the lineage level, the Mantel test revealed only a weak positive relationship between genetic and geographic distance (r = 0.063, p = 0.0004) **(Figure 6a)**.

**Figure 6:**
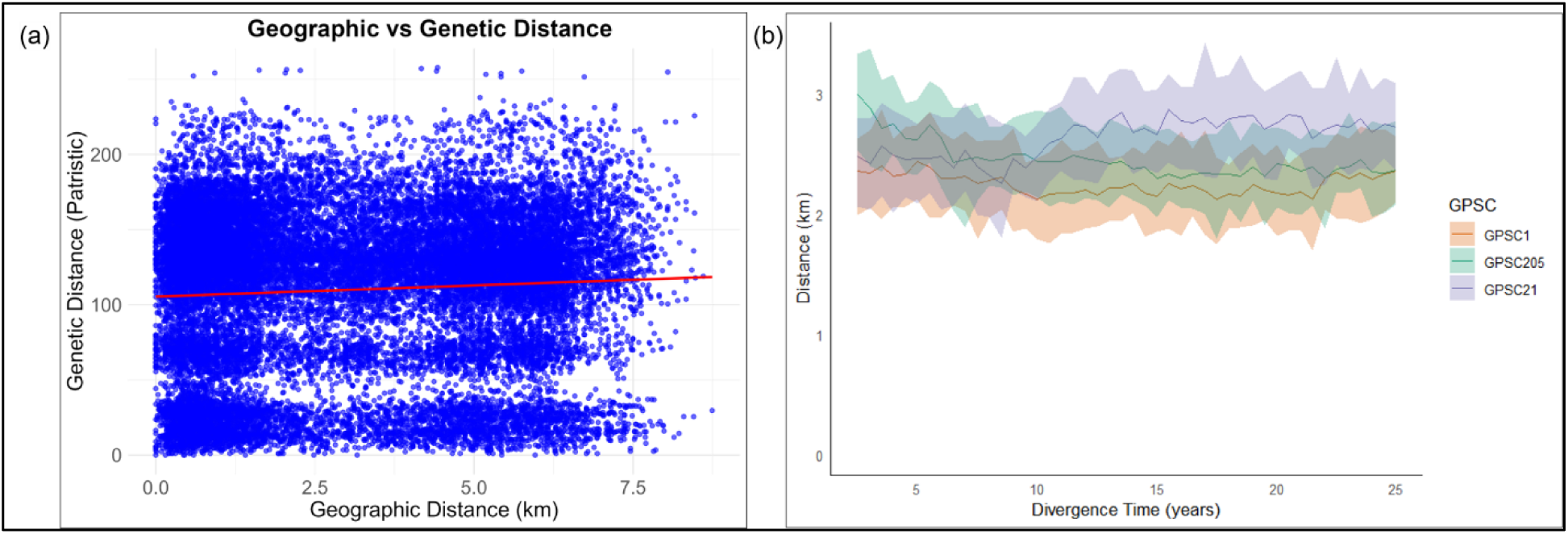
The spatial distribution and pairwise geographic distances of serotype 19F isolates. (a) The relationship between genetic and geographic distance. The red trend line indicates the correlation between spatial and genetic distances. (b) The relationship between pairwise cumulative divergence time and mean geographical distance for GPSC1 (yellow), GPSC205 (green) and GPSC21 (purple). GPSC, Global Pneumococcal Sequencing Cluster.

#### 3.4.4 Divergence time versus geographic distance

To further investigate whether genetic divergence accumulated with increasing spatial separation, we examined the relationship between divergence time and geographic distance.

Isolate pairs remained separated by approximately 2–3 km regardless of their divergence time **(Figure 6b)**, indicating little evidence that increasing evolutionary divergence corresponded to greater spatial separation.

#### 3.4.5 Spatial homogenization

Finally, to estimate how rapidly serotype 19F lineages become spatially mixed across the Drakenstein, we performed a rolling-window analysis across divergence times. Both household-level distance and regional analyses indicated rapid homogenization occurring within less than one year **(Figure S2)**. Similar results were obtained when divergence times were examined at finer temporal resolution (days).

## DISCUSSION

Persistence of pneumococcal serotype 19F despite sustained use of PCV remains one of the unresolved challenges of pneumococcal epidemiology. By integrating genomic, temporal, and spatial analyses, this study investigated the evolutionary and spatial persistence of serotype 19F in the Drakenstein, a community with high vaccine uptake. Our findings demonstrate that persistence is not driven by the expansion of a single successful clone but rather by the long-term co-circulation of multiple predominant lineages exhibiting distinct evolutionary behaviours. Although these lineages displayed localized spatial clustering, they formed a genetically well-mixed population, suggesting that the persistence is driven by the long-term co-circulation of successful lineages within a highly connected host population, where lineage-level spatial clustering persists despite extensive genetic mixing.

The evolutionary analyses revealed considerable heterogeneity among the predominant serotype 19F lineages. GPSC1 and GPSC21 exhibited measurable temporal signal over the two-decade sampling period, whereas GPSC205 showed no detectable temporal signal. These findings indicate that the predominant lineages exhibit distinct evolutionary trajectories. Such lineage-specific evolutionary dynamics are consistent with experimental studies demonstrating that bacterial populations diversify into multiple coexisting evolutionary lineages during adaptation, with different lineages exhibiting distinct phenotypic and genetic trajectories rather than a single dominant evolutionary pathway (Baym et al., 2016). Likewise, genomic surveillance in South Africa has identified GPSC1 and GPSC21 among the major serotype 19F lineages persisting following PCV introduction (Lekhuleni et al., 2024), supporting the continued success of multiple genetic backgrounds despite widespread vaccination.

Against this background, the absence of a detectable temporal signal in GPSC205 may reflect biological or epidemiological characteristics that distinguish it from the other predominant lineages. Notably, all currently available GPSC205 serotype 19F genomes in the GPS project originated from South Africa, and our analysis included all 17 available genomes. Although this may suggest a geographically restricted distribution of GPSC205, broader global sampling will be required to determine its true extent. Despite this apparent geographic restriction, GPSC205 was the predominant lineage within the Drakenstein cohort **(Table 2)**, demonstrating that local ecological success does not necessarily correspond to widespread geographic dissemination.

One possibility is that GPSC205 experiences more localized or episodic transmission than the more widely distributed GPSC1 and GPSC21 lineages, thereby reducing the accumulation of detectable temporal structure. Alternatively, differences in effective population size, recombination dynamics, or demographic history may have limited our ability to detect a temporal signal. These hypotheses remain speculative and warrant further investigation.

Although the evolutionary histories of these lineages differed, their spatial dynamics revealed a remarkably consistent pattern. Relative risk analysis demonstrated significant lineage-level spatial clustering, with isolates belonging to the same GPSC substantially more likely to occur within 1 km than expected among geographically separated isolates. Such localized clustering is consistent with the epidemiology of pneumococcal carriage, where transmission occurs predominantly through frequent close contact within households, neighbouring communities, childcare environments, and other repeated social interactions (Melegaro et al., 2004; Zhou et al., 2021; Zivich et al., 2018).

Modelling studies have further shown that pneumococcal transmission is strongly structured by age-specific contact patterns, with toddlers and older children acting as the principal drivers of transmission both within households and in the wider community, while transmission within households frequently occurs from older to younger siblings (Althouse et al., 2017). Given that the Drakenstein cohort comprised children under five years of age living predominantly in small- to medium-sized households, these contact structures likely facilitated repeated opportunities for local transmission, thereby promoting the spatial clustering of successful lineages. Together, these findings suggest that successful lineages become established within local transmission networks before gradually dispersing throughout the wider community.

However, the lineage-level clustering identified by the relative risk analysis was not accompanied by equivalent clustering of genetically related isolates. The Mantel test showed only a weak relationship between genetic and geographic distance, while the divergence time analysis demonstrated that isolate pairs remained separated by similar geographic distances regardless of the time since their most recent common ancestor. These findings suggest that although successful lineages occupy similar geographical neighbourhoods, genetically related isolates are not restricted to localized transmission foci but instead become widely dispersed throughout the study population.

Similar observations have been reported in other pathogen populations. In rural Gambia, population genomic analyses of *Plasmodium falciparum* revealed little overall genetic structure, indicating continuous recombination and mixing across the parasite population. Nevertheless, parasites isolated from the same household were substantially more likely to be genetically related than those collected from different villages, highlighting the importance of local transmission despite extensive population-wide mixing (Guery et al., 2025). Likewise, Senghore et al. (2022) found that pneumococcal strain sharing was more frequent among infants living within 5 km of one another. Sustained carriage was more common among nearby infants, whereas transient strain sharing was more frequently observed between infants living farther apart. Together, these findings suggest that local transmission generates short-range clustering, while repeated movement and transmission events continually redistribute successful genetic backgrounds across the wider population.

Taken together, our findings support a model in which neighbourhoods become enriched for successful pneumococcal lineages through repeated introductions and overlapping transmission events rather than sustained expansion of a single clone. Consequently, lineage identity retains measurable spatial structure, whereas fine-scale genetic relatedness is progressively eroded by continuous circulation within a highly connected host population.

This interpretation was further supported by the homogenization analysis. We found that serotype 19F appeared to become spatially homogenized within less than one year, even after increasing the temporal resolution to individual days. This contrasts sharply with previous studies using national surveillance datasets, where pneumococcal populations required approximately five years in Israel and nearly fifty years in South Africa to become spatially homogenized (Belman et al., 2024; Cheng et al., 2024). Rather than indicating exceptionally rapid bacterial spread, we believe the apparent early homogenization observed here reflects the characteristics of the study population and sampling framework.

The DCHS was conducted within two neighbouring peri-urban communities separated by approximately 2 km and encompassing an overall study radius of approximately 8 km. Combined with intensive longitudinal sampling, this relatively small geographical area likely captured a pneumococcal population that was already well mixed before sampling commenced. Consequently, the homogenization analysis may reflect a stable endemic transmission equilibrium rather than the active process through which lineages become geographically dispersed.

These findings suggest that persistence of serotype 19F in the Drakenstein community is maintained through continual circulation within a densely connected host population. Frequent interpersonal contact, overlapping household and neighbourhood networks, repeated exposure among young children, and prolonged asymptomatic carriage likely provide numerous opportunities for transmission and recolonization (Funk et al., 2024; Manenzhe et al., 2019; Ozella et al., 2018). Under such conditions, localized lineage clusters can repeatedly emerge through short-range transmission while ongoing host movement and repeated transmission events continually redistribute genetically related isolates throughout the community (Guery et al., 2025). This dynamic provides a plausible explanation for the coexistence of significant lineage-level spatial clustering alongside weak fine-scale genetic structure.

Interestingly, this interpretation differs from observations reported in Blantyre, Malawi, where genetically related pneumococcal isolates became progressively more geographically separated with increasing divergence time (Cave et al., 2026). Although both study settings are characterised by high carriage prevalence and dense populations, the Drakenstein cohort represents a much smaller and more geographically constrained community. These differences highlight how spatial scale, population structure, and sampling design can substantially influence the inferred dynamics of pneumococcal spread, even among epidemiologically similar settings.

Several limitations should be considered when interpreting these findings. First, our analyses focused exclusively on serotype 19F, whereas natural pneumococcal carriage frequently involves sequential or concurrent colonisation by multiple serotypes and lineages (Almeida et al., 2026; Gladstone et al., 2020). Interactions among co-colonising strains, together with horizontal gene transfer and within-host evolution, may influence transmission dynamics in ways not captured by single-serotype analyses (Chaguza et al., 2020; Sibale et al., 2025). Second, the study was conducted within a relatively small geographical area, which may limit extrapolation of these findings to larger regional or national populations. Finally, although the incorporation of the GPS dataset substantially strengthened the evolutionary analyses, differences in sampling strategies across contributing studies may have influenced estimates of temporal signal and evolutionary history.

Overall, our findings demonstrate that persistence of pneumococcal serotype 19F cannot be explained solely by changes in serotype prevalence or the expansion of a single dominant clone. Instead, persistence appears to result from the long-term circulation of multiple successful lineages within a highly connected host population, where lineage-level spatial clustering is continually counterbalanced by rapid genetic mixing. Here, we show that understanding why VTs persist requires not only identifying which lineages are circulating but also understanding how they evolve and spread through local host populations.

By integrating evolutionary analyses with fine-scale spatial epidemiology, this study provides new insights into the mechanisms underlying the persistence of pneumococcal serotype 19F in a highly vaccinated population. While previous genomic studies have successfully characterised pneumococcal spread across broader geographic scales, our findings demonstrate the value of applying these approaches at the micro-community level, where transmission dynamics may differ substantially. By jointly examining lineage evolution, spatial clustering, genetic mixing, and temporal dynamics, we show that persistence reflects the interaction between evolutionary processes and local transmission dynamics within a densely connected host population. These findings highlight the importance of integrating genomic surveillance with spatial epidemiology to better understand pathogen persistence and may help inform future surveillance strategies and public health interventions in high-burden settings.

## Data Availability

All data produced in the present study are available upon reasonable request to the authors.

## APPENDICES

**Figure S1.**
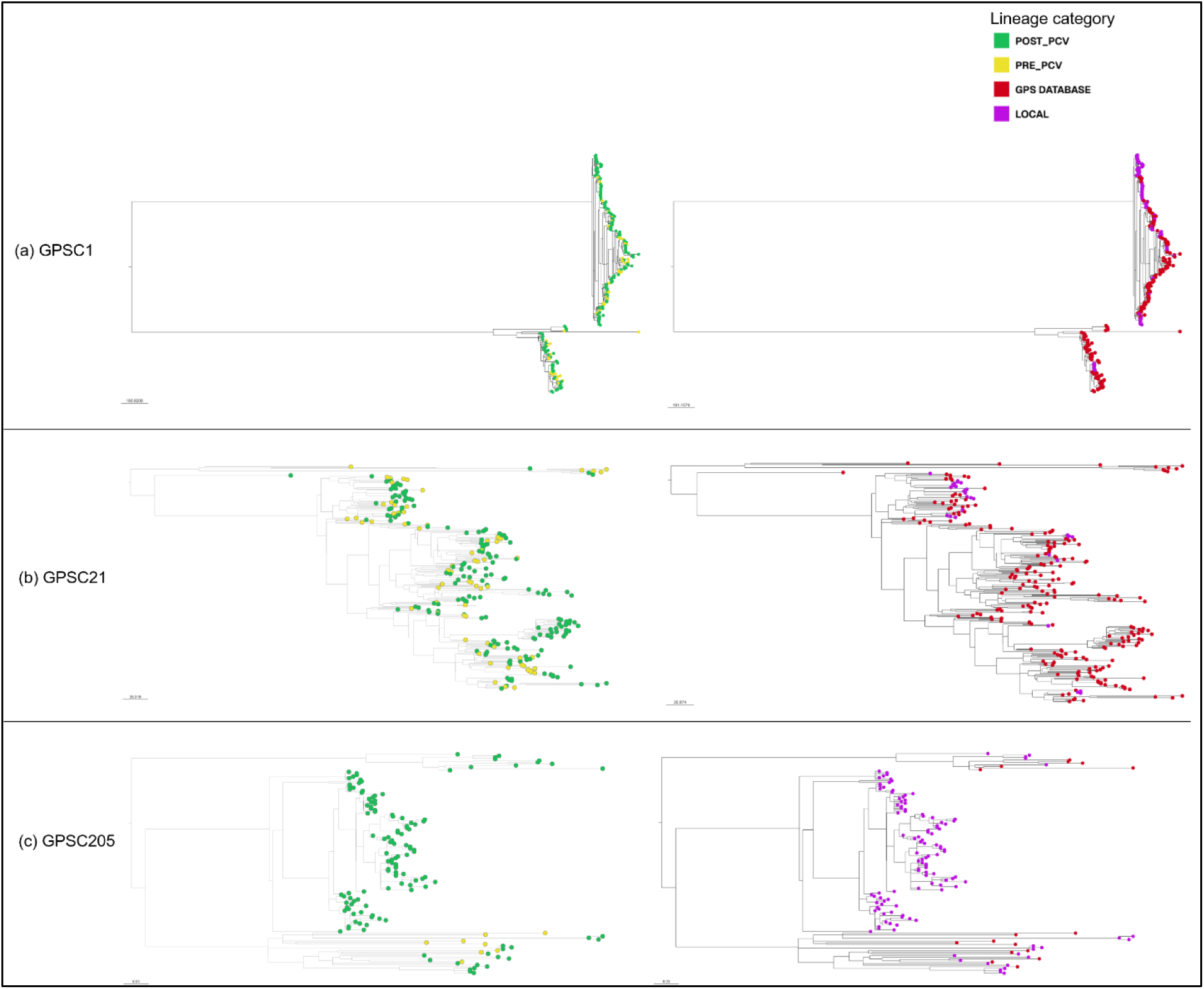
Time-scaled phylogenetic relationships of serotype 19F lineages. Maximum clade credibility trees for GPSC1 (top), GPSC21 (middle), and GPSC205 (bottom) inferred using time-scaled phylogenetic analyses. Trees in the left panels are coloured according to vaccine era, with isolates collected during the pre-PCV period shown in yellow and those collected during the post-PCV period shown in green. Trees in the right panels are coloured according to isolate source, with local Drakenstein isolates shown in purple and isolates obtained from the Global Pneumococcal Sequencing (GPS) database shown in red. PCV, pneumococcal conjugate vaccine; GPSC, global pneumococcal sequencing cluster.

**Figure S2.**
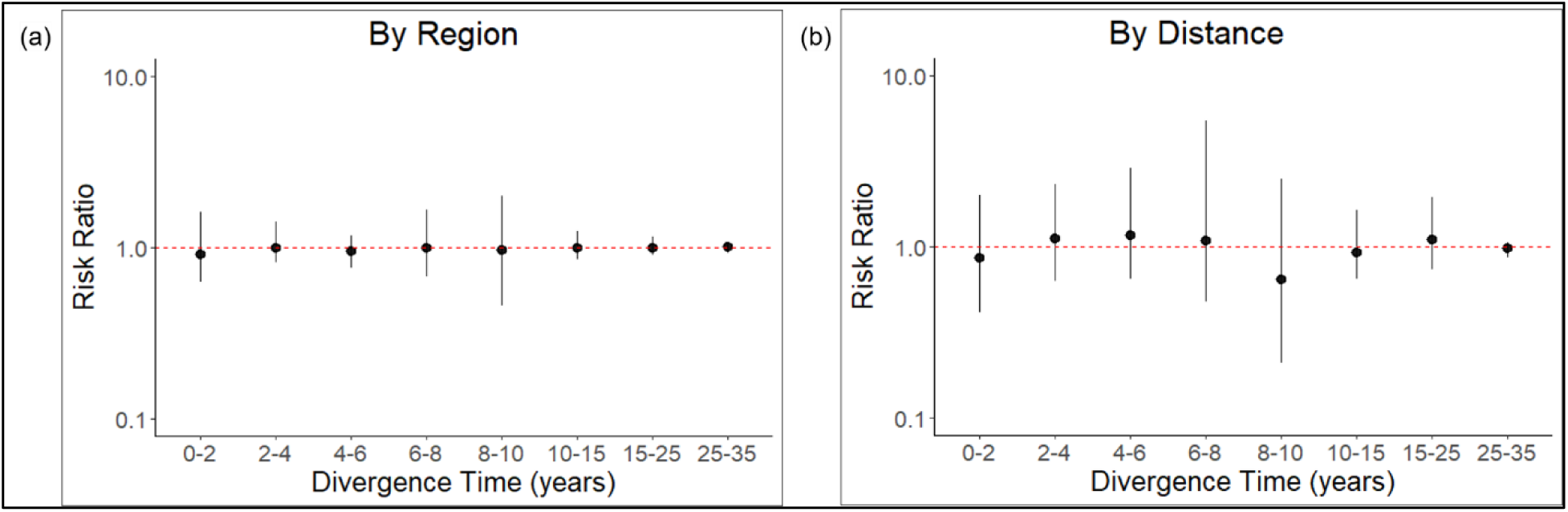
Spatial homogenization of pneumococcal serotype 19F lineages. The relationship between the pairwise divergence time and the relative risk ratio for the dominant serotype 19F lineages in the Drakenstein when comparing within and between regions (Mbekweni and TC Newman) on the left and when comparing pairs of isolates within 2 km and pairs of isolates larger than 5 km apart between distance on the right. The red dashed line indicates a relative risk of 1 (no difference in risk). Solid black dots and lines represent the 2.5, 50, and 97.5 percentiles of the confidence intervals.

